# Longitudinal Influenza A Virus Screening of Retail Milk from Canadian Provinces (Rolling Updates)

**DOI:** 10.1101/2024.05.28.24308052

**Authors:** Hannah L. Wallace, Jordan Wight, Mariana Baz, Barbara Dowding, Louis Flamand, Tom Hobman, Francois Jean, Jeffrey B Joy, Andrew S. Lang, Sonya MacParland, Craig McCormick, Ryan Noyce, Rodney S. Russell, Selena M. Sagan, Jumari Snyman, Gabriela J. Rzeszutek, Mustafa S. Jafri, Isaac Bogoch, Jason Kindrachuk, Angela L. Rasmussen, the Pan-Canadian Milk Study Network

## Abstract

Highly pathogenic avian influenza (HPAI) H5N1 has caused the deaths of more than 100 million birds since 2021, and human cases since 1997 have been associated with significant morbidity and mortality. Given the recent detection of HPAI H5N1 in dairy cattle and H5N1 RNA detections in pasteurized retail milk in the United States, we established the Pan-Canadian Milk (PCM) Network. Through our network of collaborators from across Canada, retail milk is being procured longitudinally and sent to a central laboratory for testing for the presence of influenza A virus RNA. To date (05 July 2024), we have tested 92 retail milk samples from all ten Canadian provinces (NL, NS, PEI, NB, QC, ON, MB, SK, AB, and BC) and all have tested negative for influenza A virus RNA. Testing is ongoing and these results will be updated on rolling basis as additional data becomes available. Despite no known HPAI infections of dairy cattle in Canada to date, H5N1 poses a significant threat to the health of both humans and other animals. Routine surveillance of retail milk on a national scale will allow for monitoring of infected dairy cattle on an ongoing basis in a cost-effective, standardized, scalable and easily accessible manner. Our network and testing will act as an early detection system to help inform containment and mitigation activities if positive samples are identified.

## Introduction

Influenza A viruses (IAVs) are in the family *Orthomyxoviridae* which can infect a wide variety of hosts and result in a wide spectrum of illness, from asymptomatic to severe including multi-organ failure resulting in death (1, 2). Highly pathogenic avian influenza (HPAI) is an emerging disease threat with pandemic potential. Most recently arriving in North America in late 2021 (3), HPAI H5N1 clade 2.3.4.4b has infected a broad range of wild and domesticated bird species, and has resulted in the death of more than 100 million birds (4). This virus and derived reassortants have also infected diverse species of marine and terrestrial mammals (5–8), with the most recent infections occurring in dairy cattle in the United States (9–11). Since 1997, spillover of HPAI H5N1 from birds to humans have also been documented. Of the nearly 900 known human infections of H5N1, most have exhibited severe disease symptoms, with a case fatality rate of approximately 50% (12). However, despite the fact that the currently circulating H5N1 clade 2.3.4.4b virus and derived reassortants have caused extensive infections and deaths of birds and mammals, relatively few human cases have been reported, with only 13 cases globally associated with this clade since January 2022 (12, 13). Notably, none of these cases have been fatal, with the majority being reported as exhibiting mild disease symptoms with frequent reports of conjunctivitis. Four of the 12 human cases were reported in the spring/summer of 2024; cases were associated with close interactions with infected dairy cattle. The four cases all had relatively mild symptoms (conjunctivitis), with one also reporting mild respiratory symptoms and all recovered (14–17). In response to the first human case associated with dairy cattle in Texas, the CDC experimentally infected ferrets with the human isolate, A/Texas/37/2024(H5N1). In ferrets, the isolate from the individual who was infected in Texas, was able to infect ferrets efficiently and caused 100% lethal disease. Interestingly, unlike transmission of seasonal IAVs in ferrets, the Texas isolate was only able to spread ferret-to-ferret through direct contact and not through respiratory droplets (18).

The United States Department of Agriculture (USDA) reported in late March 2024 that dairy cattle on a farm in Texas were exhibiting signs of lethargy, dehydration, mild respiratory symptoms, decreased feed intake, decreased milk production and/or milk with abnormal colour/texture, and further investigation revealed lesions similar to those seen with mastitis (19, 20). There were also reports of barn cats being found dead on this property (20). Phylogenetic analyses showed that the infection of dairy cattle was the result of a single spillover event of HPAI H5N1 from wild birds in late 2023 (21). This was then followed by subsequent cattle-to-cattle transmission possibly mediated by exposure to contaminated milking equipment, or another currently unknown route (10, 22). Since then, the outbreak has spread to 140 herds in the US as the transport of infected dairy cattle occurred before the extent of the outbreak was known (23). H5N1-infected dairy cattle have now been reported in 12 states (23) and recently H5N1 RNA was detected in wastewater from nine cities across Texas (24).

Soon after the initial dairy infections were reported, the FDA began conducting H5N1 surveillance of pasteurized milk and other dairy products from 38 states and reported that up to 20% of samples tested were positive for H5N1 RNA. Importantly, no viable virus was detected in the retail pasteurized dairy samples (25). The USDA has also undertaken testing of raw milk (n=275) from states known to have H5N1-infected herds and found that 158 (58%) of the samples were positive for viral RNA with 39 (25%) of the RNA positive samples also being containing infectious virus. Despite the high proportion of positive samples, the group also that continuous flow pasteurization (commonly used and FDA approved) of the milk completely inactivated any infectious virus, indicating that the milk supply is safe despite the detection of viral RNA (26). Testing of tissue from dairy cattle as well as testing of retail beef is also being performed by the USDA. To date, one muscle sample from a dairy cow known to be infected with H5N1 has shown to be positive for H5N1 RNA and no meat from any dairy cattle has entered the US food supply (27).

To date, no H5N1 infections of dairy cattle have been reported in Canada. Due to current mandatory testing by the USDA before transporting cattle across state boarders to limit inter-state spread (28), along with limited importation of dairy cattle from the US into Canada, infected cattle are unlikely to enter Canada and threaten Canadian cattle operations. Additionally, since 29 April 2024, the Canadian Food Inspection Agency (CFIA) has implemented a requirement for proof of negative testing on USDA export certificates for all lactating dairy cattle (29), further protecting Canadian cattle. Furthermore, as this virus still extensively circulates in wild birds, there is the potential for the virus to independently spill over from wild birds into Canadian cattle, as occurred on the index farm in Texas. Therefore, monitoring of cattle in Canada is of vital importance to detect if a potential spillover has occurred as quickly as possible.

Although it is known that cattle can be infected by influenza A viruses (30–32), including H5N1 (33), no viral genomes have been sequenced from cattle, prior to early 2024. Very little is known about the patterns of disease or the tissue types that allow viral replication although decreased milk production had been previously associated with influenza antibodies in dairy cattle (34). A recent pre-print suggested that mammary tissue of dairy cattle express both α-2,3- and α-2,6-sialic acid, associated with avian and human influenza receptors, respectively (35). This suggests the possibility of dairy cattle acting as a “mixing pot” for avian and human influenza virus reassortment, similar to what has been described for swine (10). Additionally, Bordes *et al*. recently found that H5N1 viruses from the same clade as the dairy cattle viruses (2.3.4.4.b) that were originally isolated from a chicken and a fox in the Netherlands were able to efficiently replicate in well-differentiated bovine airway epithelial cells cultured at an air-liquid interface (36). Additionally, scientists in Germany recently reported that an H5N1 virus isolated from a wild bird was able to infect the udder of experimentally infected cattle (37). These findings suggest that other H5N1 viruses may also have the potential to infect cattle, highlighting the need for ongoing surveillance (36, 37).

In April 2024, we established the Pan-Canadian Milk (PCM) Network, bringing together colleagues from across the country to procure retail milk every two weeks and send samples to a central laboratory for testing. This study will continue longitudinally, with particular interest in testing during the wild bird migration seasons. Recently, the CFIA as well as academic colleagues in the province of Ontario tested retail milk and found that all samples tested IAV RNA negative; however, these efforts represent a snapshot in time and no detailed plans for longitudinal screening have been announced by either group (38, 39). Ongoing surveillance in retail milk will not only allow rapid containment measures should any samples test positive but may also further our understanding of dairy cattle susceptibility to various influenza A viruses, not just H5N1.

## Methods

### Milk Sampling

Pasteurized whole (3.25%) milk was obtained from local retailers by collaborators in all Canadian provinces. However, additional detailed data was collected about each retail milk sample though this is not reported here. Samples were collected every two weeks beginning 29 April 2024. Milk cartons were externally disinfected, opened in a biosafety cabinet, and a sample aseptically collected in sterile conical tubes at the laboratories of collaborators. Samples were kept at 4°C and shipped on ice to the Kindrachuk lab at the University of Manitoba for processing. Samples were then stored at −80°C.

### RNA Isolation

RNA isolation was performed from 140 µL of milk using the Qiagen Viral RNA Mini Kit (Qiagen, Product # 52906) as per the manufacturer’s instructions. Isolated RNA was stored at −80°C.

### Screening for Influenza A Virus

Real-time RT-PCR was used to screen for the presence of IAV matrix gene RNA as per Wight *et al*. (40) using a QuantStudio 6 Flex instrument (Applied Biosystems). Any sample that tests positive for the matrix gene will be subsequently screened for the H5 subtype of the haemagglutinin gene also as per Wight *et al*. (40). In the event of positive sample identification, data will be immediately shared with federal agencies through existing linkages and collaborative connections.

## Results (Rolling Updates)

As of 05 July 2024, 92 retail milk samples obtained from each of the Canadian Provinces (NL, NS, PEI, NB, QC, ON, MB, SK, AB, and BC) have been tested. All samples to date have tested negative for IAV matrix RNA (Table 1).

**Table 1.** Provinces and pan-Canadian totals of retail milk samples tested, the number of IAV RNA positive samples, and the percentage of IAV RNA positive samples. RNA was isolated from milk samples and screened for IAV matrix RNA using real-time RT-PCR.

| Province | Number of Milk Samples Tested | Number of IAV RNA Positive Samples | % of IAV RNA Positive Samples |
| --- | --- | --- | --- |
| Newfoundland and Labrador | 8 | 0 | 0% |
| Nova Scotia | 6 | 0 | 0% |
| Prince Edward Island | 1 | 0 | 0% |
| New Brunswick | 8 | 0 | 0% |
| Quebec | 15 | 0 | 0% |
| Ontario | 7 | 0 | 0% |
| Manitoba | 9 | 0 | 0% |
| Saskatchewan | 13 | 0 | 0% |
| Alberta | 14 | 0 | 0% |
| British Columbia | 11 | 0 | 0% |
| **National Totals** | **92** | **0** | **0%** |

## Discussion

CFIA has recently reported that the 600 milk samples they tested were H5 RNA negative (38) as well as the fact that no outbreaks of disease associated with H5N1 have been reported in dairy cattle in Canada, it was not unexpected that our samples, collected during nearly the same time as CFIA’s, also tested negative for influenza.

Given the expansive nature of the H5N1 outbreak in dairy cattle, spillover to other species, and at least four human infections linked to close contact with dairy cattle in the US, continued longitudinal monitoring of milk samples in Canada may allow early detection of HPAI infection of dairy cattle. This will augment and compliment ongoing efforts by government agencies and other academic groups, enabling early containment and preventative measures to inhibit further spread. Sample collection and testing will be ongoing for the foreseeable future. This document will be updated as additional data becomes available. Our results will be provided to federal government agencies in real-time as this live preprint is updated to ensure early, transparent, and collaborative data exchange with key stakeholders and policy makers.

## Data Availability

All data produced in the present study are available upon reasonable request to the authors

## Acknowledgements

We would like to acknowledge all PCM Study collaborators for contributing milk samples from across the country.

## Funding

This work was supported by the Canadian Institutes of Health Research (Tier 2 Canada Research Chair, grant number 950-231498 to JK) and by the Natural Sciences and Engineering Research Council Discovery Grant (RGPIN-2018-06036 to JK).

## Author Contributions

Conceptualization: HLW, JW, IB, AR, JK

Data Curation: HLW, JW

Formal Analysis: HLW, JW

Funding Acquisition: JK

Investigation: JW, HLW

Methodology: JW, HLW

Project Administration: HLW, JW, JK

Resources: JK

Supervision: JK

Writing (original draft): HLW, JW

Writing (review and editing): All authors

## Competing Interests

The authors declare that they have no competing interests.

## Notes

### Competing Interest Statement

The authors have declared no competing interest.

### Summary of Updates

This revision includes our latest testing updates as of July 5, 2024

